# High prevalence of deleterious germline variants in cancer risk genes among subjects with young-onset, sporadic pituitary macroadenomas

**DOI:** 10.1101/2025.05.15.25327006

**Authors:** Adrian F. Daly, Kalyani Sridharan, Marie-Lise Jaffrain-Rea, Giampaolo Trivellin, Francesca Carbonara, Wouter De Herder, Ismene Bilbao, Maria Segni, Margaret Zacharin, Mariana Solovey, Kavita Kadian, Ulrich Paetow, Nalini Shah, Tushar Bandgar, Liliya Rostomyan, Sebastian J.C.M.M. Neggers, Albert Beckers, Patrick Pétrossians

**Author notes:** Correspondence should be addressed to: Adrian F. Daly MD, PhD, Department of Endocrinology, Centre Hospitalier Universitaire de Liège (B35), University of Liège, Domaine Universitaire Sart-Tilman, 4000 Liège, Belgium. **Disclosure**: The authors have nothing to disclose that would impact the presentation or interpretation of the study.

## Abstract

**Introduction:** Pituitary adenomas/pituitary neuroendocrine tumors (PitNETs) are common intracranial tumors, clinically affecting 1:1000 individuals and most cases remain genetically unexplained. Emerging research has highlighted the major contribution of germline pathogenic variants to tumorigenesis across many tissue types in young subjects . We investigated whether young-onset (<30 years old) pituitary macroadenomas that were negative for known genetic causes harbor pathogenic or likely pathogenic (P/LP) variants in cancer-risk genes.

**Methods:** We retrospectively analyzed 48 subjects (29 males; 96% GH- or PRL-secreting) with sporadic pituitary macroadenomas that were negative for known germline variants (*AIP, MEN1, CDKN1B*) or duplications (*GPR101*). Whole-exome sequencing (WES) was performed on germline DNA. Bioinformatics analysis including variant calling (for small variants and CNVs), annotation and variant prioritization were performed, using secondary analysis pipelines for WES data and AION predictor platform for tertiary analysis. Variants in established cancer-risk genes were prioritized.

**Results:** P/LP germline variants in cancer-risk genes were identified in 14.6% of subjects on ClinVar/ACMG criteria. This rose to 31.3% of subject with deleterious variants when additional *in silico* and AION predictor criteria were used. Genes included: *BAP1, BRCA1, BUB1, ELAC2, FLCN, MCPH1, MSR1, MUTYH, PDE11A, POLE, POLG, PMS2, RAD51C, RECQL4, SDHA, SDHD, SEC23B, TMEM127, WRN*.

**Conclusions:** Our findings expand the spectrum of genes potentially associated with young-onset pituitary macroadenomas. The identification of a high rate of deleterious germline variants in cancer-risk genes in pituitary adenomas/PitNETs echoes similar findings in young patients across a wide range of tumors. These results may have relevance for genetic counseling and potentially could expand targeted management strategies in young patients with large pituitary tumors.

## Introduction

The anterior pituitary gland is a neuroendocrine organ that plays a crucial role in regulating many of the body’s major functions. Pituitary adenomas (termed pituitary neuroendocrine tumors (PitNETs)) occur incidentally in about 20% of the population, whereas clinically-relevant pituitary tumors occur in around 1:1000 of the general population (1–4). Pituitary tumors are also the most frequent primary brain/central nervous system tumors in adolescents and young adults (5,6). Although these tumors very rarely metastasize, some are aggressive and locally invasive, impinging on vital brain structures like cranial nerves and carotid vessels (7). The combination of local tumoral effects and/or hormonal dysregulation leads to classical endocrine diseases like acromegaly-gigantism, prolactinoma, Cushing’s disease and clinically non-functioning pituitary adenomas (7).

The pathophysiology of pituitary adenomas/PitNETs is only partially understood. At the somatic level, gain of function variants account for up to 40% of cases, such as *GNAS* in acromegaly (8,9) and *USP8* in Cushing’s disease (10,11). Other recurrent somatic variants are relatively uncommon, or are still emerging (e.g., *SF3B1* variants in prolactinomas) (12–14). Comprehensive genomic studies have classified tumor tissue by cell lineage and chromosomal instability, which can predict aspects of clinical behavior (3,15–21). Germline genetic causes are infrequent and explain about 5% of pituitary adenomas overall (22). Among these, the most prevalent are rare loss of function (LOF) variants in *AIP* that cause young onset and familial isolated pituitary adenomas (FIPA), primarily acro-gigantism and prolactinomas (23–26). Duplications disrupting a conserved topologically associating domain (TAD) at *GPR101* cause sporadic and familial X-linked acrogigantism (X-LAG) (27–30). Pituitary adenomas also form part of well-characterized syndromes like multiple endocrine neoplasia (MEN) 1, MEN4 and Carney complex due to LOF germline variants in *MEN1*, *CDKN1B*, and *PRKAR1A*, respectively (31–33). Pituitary adenomas are rare clinical manifestations of other genetic disorders, such as, pheochromocytoma-paraganglioma syndromes (34–37).

Recently, genetic research in oncology has shifted focus on to the role of deleterious germline genetic variants in the etiologies of various tumor types. Elevated rates of germline pathogenic/likely pathogenic (P/LP) variants in cancer-risk genes have been reported across a wide spectrum of neoplasias (38–40). Such findings have been reported in sporadic thyroid cancer and pancreatic neuroendocrine tumors (41–45). Some studies using limited gene panels suggest that germline P/LP variants in cancer-risk genes might also occur in pituitary tumors (46–53). Identifying novel causes has direct clinical implications for family screening, and for personalized molecular therapies (54). To test the hypothesis that cancer-risk genes could be involved in pituitary tumorigenesis in “high-risk” populations, we performed an analysis of germline whole exome sequencing (WES) data using a comprehensive cancer gene panel in a cohort of subjects with young-onset pituitary macroadenomas in whom established germline genetic causes of pituitary tumors had been ruled out.

## Methods

### Subjects

Subjects belonged to a retrospective, international study on the genetic causes of pituitary adenomas. To be included in the current analysis, subjects had to have an isolated pituitary macroadenoma (≥10 mm maximum diameter) on imaging that was diagnosed before 30 years of age in the absence of FIPA. In addition, using next generation sequencing (NGS) or Sanger sequencing, all subjects had to be negative for pathogenic germline variants in genes associated with early-onset pituitary adenomas or FIPA (*AIP*, *MEN1*, and *CDKN1B*). Copy number variants in those genes were ruled out using MLPA in all patients. Subjects with pediatric-onset gigantism (n=14) had to be negative on array comparative genome hybridization studies (aCGH) for duplications involving *GPR101* on chromosome Xq26.3.

Demographic, clinical, and hormonal data were collected. General data on a personal family history of cancer or other medical conditions were not collected systematically.

The work was approved by the Ethics Committee of the CHU de Liège-University of Liège under study codes GENOCRINE-2021/213, B70720109577, B707201420418, and B707201111968. All subjects or their guardians provided written informed consent in their own language.

### Genetic analyses

Germline analyses were performed using leukocyte-derived DNA. Samples were pre-checked for DNA amount and concentration using a fluorescence-based method, Qubit dsDNA BR (Thermo Fisher). Libraries were prepared using 50 ng (ultra-low input), 200 ng (low input) and 1 µg (standard input) of DNA and a HiSeq platform with 2×100 bp was used. Sequencing reads were demultiplexed with Illumina bcl2fastq (2.19 or 2.20). Adapters were trimmed with Skewer (version 0.2.2) (55). Reads were mapped to the reference genome hg19. The qualities of the FASTQ files were analyzed with FastQC (version 0.11.5-cegat). Plots were created using ggplot2 in R (version 4.0.4) (R Core Team) (56,57).

### Bioinformatic pipeline

WES data was analyzed by Nostos Genomics for secondary and tertiary analysis. Small Variant detection (SNPs and Indels) was carried out using Sentieon DNAScope 5.0.1. Tertiary analysis for variant annotation and interpretation was performed in the AI-powered platform AION predictor (Nostos Genomics GmbH, Berlin, Germany). The general workflow for this AI-assisted algorithmic platform for genetic variant interpretation is outlined below and in Supplemental Materials. Vcf files obtained from the secondary analysis were submitted to the AION predictor tertiary analysis pipeline that starts with data preprocessing. The primary goal of this step is to read and prepare the input VCF files for subsequent annotation and analysis. Thereafter, the data were annotated to add genetic, molecular and clinical information (e.g., gene involved, prevalence in the population, known disease associations). These annotated variants were then classified by their potential to cause disease using ClinVar classifications, automated implementation of ACMG guidelines and the proprietary AION predictor that integrates comprehensive data from external databases covering epidemiological, molecular and other software prediction sources. Concurrently, the patient’s symptoms and physical findings were cataloged using Human Phenotype Ontology (HPO) terms. A comprehensive soft filter was applied to focus the analysis on genes associated with a hereditary propensity to solid tumors, including both endocrine and non-endocrine tissues (Supplemental materials).

In the case of missense variants in any of the reported genes, we also applied the AlphaMissense tool to assess the potential pathogenic or benign nature. AlphaMissense is an AI prediction tool that is based on AlphaFold technology (Google Deepmind) that is trained on population frequency data, protein language modeling of amino acid distribution in proteins and protein structure context (58,59). AlphaMissense does not utilize annotations from clinical/human sources. AlphaMissense assigns a score to predict whether a missense variant is classified as likely benign, likely pathogenic, or uncertain (59). For splice-altering variants, we included analyses based on the SpliceAI tool and a new heuristic model for splice altering variant (SAV) interpretation described by Sullivan *et al* (20,60). The SpliceAI tool is based on a 32-layer deep neural network that predicts splicing from pre-mRNA sequences. For SpliceAI, we included splice variants with a score of >0.8 to assign a deleterious effect. In the heuristic model only variants with a high SAV assessment (>90%) were retained.

Finally, variants were tabulated, presented and discussed under three separate groupings: (1) variants with P/LP classifications from ACMG and/or Clinvar, including supporting pathogenicity classifications on AION predictor and other models; (2) variant with VUS, conflicting or uncertain significance classifications on ACMG/CLinvar, but with P/LP scores on AION predictor and other models; (3) gene variants with ambiguous classifications on models and/or high population prevalences.

### Copy number variation analysis

The ExomeDepth pipeline was used as a computational method to detect copy number variants (CNVs) from exome sequencing data using read depth information (61). The input files needed are BAM files (aligned sequencing reads) BED files, to define the targeted region, and a reference cohort with samples from unrelated, unaffected individuals that were processed using the same capture kit during library preparation and the same sequencing platform. For the sample CNV calling ExomeDepth selects the best-matched reference samples to minimize noise and to help correct for biases that might arise due to the inconsistent capture efficiency across exons or targeted regions. For tertiary analysis of the derived data, the following variant criteria were used to filter and report variants from the cancer gene panel listing: pathogenic (P) and likely pathogenic (LP) variants by

ACMG classification; VUS (Variant of Uncertain Significance) variants by ACMG classification associated with a known disease gene; variants with gnomAD frequency ≤ 1%, if gnomAD data were available. The included deletions and duplications were sorted based on ACMG classification, phenotypic score and ACMG score, ensuring the most clinically relevant and evidence-supported variants were prioritized at the top of the relevant list.

### Pathway analysis

We performed an enrichment analysis to interrogate genes identified in this study. Metascape is a validated system that includes inputs from a wide range of databases and incorporates computational analysis pipelines that are regularly updated and synchronized (62,63) . Metascape permits gene annotation, membership analyses, and meta-analyses. For the current study we visualized gene enrichment using Gene Ontogeny/KEGG terms and the DisGeNET database, which were outputted as bar charts. For statistical significance, the cut-off for the expression analysis was taken as *p* <2 × 10^−6^.

## Results

### Study population

We studied 48 subjects with isolated, sporadic pituitary macroadenomas that were diagnosed before 30 years of age (60.4% males). Demographic and clinical data are shown in Table 1. The median age at diagnosis overall was 18 years (range: 3-30 years). Among the group, 10 were diagnosed aged ≤12, 20 were aged from 13-20 years and 18 were 21-30 years of age at diagnosis (52.1% children/adolescents). There were 35 individuals with GH-secreting pituitary tumors of whom 14 met height/growth criteria for pituitary gigantism (64).

**Table 1.** Clinical and demographic details on subjects with young-onset pituitary macroadenomas. Age ranges: childhood (0-12 years); adolescent (13-18 years); young adult (19-30 years). F: female; GH: growth hormone secreting; M: male; Macro: macroadenoma; NFPA: non-functioning pituitary adenoma; PRL: prolactinoma.

| **Subject Number** | **Sex** | **Pituitary Adenoma Type** | **Age at Onset (range)** | **Tumor classification** | **Max tumor diameter (mm)** |
| --- | --- | --- | --- | --- | --- |
| 1 | M | GH | childhood | Macro | 20 |
| 2 | M | GH (acrogigantism) | adolescent | Macro | N/A |
| 3 | M | GH (acrogigantism) | childhood | Macro | 33 |
| 4 | M | GH (acrogigantism) | childhood | Macro | N/A |
| 5 | F | GH | childhood | Macro | N/A |
| 6 | M | GH (acrogigantism) | childhood | Macro | N/A |
| 7 | M | GH (acrogigantism) | childhood | Macro | 27 |
| 8 | M | GH (acrogigantism) | adolescent | Macro | 30 |
| 9 | M | GH (acrogigantism) | adolescent | Macro | N/A |
| 10 | M | GH (acrogigantism) | adolescent | Macro | N/A |
| 11 | F | GH | young adult | Macro | 40 |
| 12 | M | PRL | childhood | Macro | 70 |
| 13 | M | GH (acrogigantism) | adolescent | Macro | 60 |
| 14 | F | GH | childhood | Macro | N/A |
| 15 | M | GH (acrogigantism) | adolescent | Macro | N/A |
| 16 | F | GH (acrogigantism) | adolescent | Macro | N/A |
| 17 | F | PRL | childhood | Macro | N/A |
| 18 | F | GH (acrogigantism) | childhood | Macro | N/A |
| 19 | F | GH | young adult | Macro | N/A |
| 20 | M | GH | young adult | Macro | 40 |
| 21 | M | GH | young adult | Macro | N/A |
| 22 | F | PRL | young adult | Macro | N/A |
| 23 | F | PRL | young adult | Macro | N/A |
| 24 | M | PRL | young adult | Macro | 38 |
| 25 | M | PRL | adolescent | Macro | 59 |
| 26 | F | GH | adolescent | Macro | 35 |
| 27 | F | NFPA | young adult | Macro | 29 |
| 28 | F | GH | young adult | Macro | 45 |
| 29 | F | GH | young adult | Macro | 32 |
| 30 | M | GH (acrogigantism) | adolescent | Macro | 65 |
| 31 | M | PRL | young adult | Macro | N/A |
| 32 | F | GH | young adult | Macro | 36 |
| 33 | F | GH | young adult | Macro | 37 |
| 34 | F | GH | young adult | Macro | 29 |
| 35 | M | GH | adolescent | Macro | 20 |
| 36 | M | GH | young adult | Macro | 26 |
| 37 | M | PRL | adolescent | Macro | N/A |
| 38 | F | GH | adolescent | Macro | 23 |
| 39 | M | GH | young adult | Macro | 18 |
| 40 | F | GH | young adult | Macro | 37 |
| 41 | M | GH | young adult | Macro | 40 |
| 42 | M | GH | young adult | Macro | 33 |
| 43 | F | GH | young adult | Macro | N/A |
| 44 | M | PRL | adolescent | Macro | 35 |
| 45 | M | PRL | young adult | Macro | 48 |
| 46 | M | PRL | young adult | Macro | N/A |
| 47 | M | NFPA | adolescent | Macro | N/A |
| 48 | M | GH<br>(acrogigantism) | young adult | Macro | N/A |

### Genetic analysis and variant identification

There were 7/48 (14.6%) subjects with heterozygous P/LP germline variants on ACMG and/or ClinVar criteria (Table 2). Another 11 variants in 10 genes were classified as VUS or conflicting interpretation on ACMG/ClinVar: one was the loss of the start codon, one was an early truncating variant, six were missense variants, and two were splice variants. All missense variants had pathogenic scores on AlphaMissense and the two splice variants were scored as pathogenic by SpliceAI/SAV. Finally, two missense variants in *SDHA* and *TSC2* and a relatively prevalent *PDE11A* truncating variant were classed as VUS/conflicting interpretation on ACMG/ClinVar; all three were P/LP on AION predictor but the missense variants were ranked as ambiguous by AlphaMissense and these three variants were not included in the overall calculations. Analysis of the variants using the AION predictor scored 15/48 (31.3%) of subjects as having a deleterious variant.

**Table 2.** Germline variants of interest in cancer-related genes identified during WES analysis in 48 young subjects with previously genetically negative pituitary adenomas. Age ranges: childhood (0-12 years); adolescent (13-18 years); young adult (19-30 years). AION: AION Predictor; AM: Alpha missense; CI: conflicting interpretations; LB: likely benign; LP: likely pathogenic; P: pathogenic; PA: pituitary adenoma; SAV: Splicing affecting variant; US: uncertain significance; VUS: variant of unknown significance.

| Subject No. | Sex | Age at dx | PA | Gene | Nucleotide change<br>Amino acid change<br>chromosomal location (hg19) | Variant type | GnomAD Freq. | Clin Var | ACMG | AION | AM Class (Score) | SpliceAI effect (site)/ SAV heuristic (%) |
| --- | --- | --- | --- | --- | --- | --- | --- | --- | --- | --- | --- | --- |
| **P/LP: ACMG/ClinVar** |  |  |  |  |  |  |  |  |  |  |  |  |
| 8 | M | adolescent | GH | *MUTYH* | c.925-2A>G<br>-<br>1-45797760-T-C | splice acceptor | 0.0011 | CI | LP | P | - | 0.95 (AL)/<br>99.7% |
| 9 | M | adolescent | GH | *BRCA1* | c.2217_2218insA<br>p.Val740SerfsTer3<br>17-41245330-C-CT | frameshift | 0 | P | P |  |  |  |
| 11 | F | Young adult | GH | *ELAC2* | c.560-2A>G<br>17-12915101-T-C | splice acceptor | 0.00089 | CI | LP | P | - | 0.69 (AL)/<br>99.7% |
| 12 | M | childhood | PR L | *FLCN* | c.890_893del<br>p.Glu297AlafsTer25<br>17-17122501-GCTTT-G | frameshift | 0.00001 | P | P |  |  |  |
| 15 | M | adolescent | GH | *RECQL4* | c.2398C>T<br>p.Gln800Ter<br>8-145738666-G-A | stop gained | 0.00001 | P | LP | P | - |  |
| 18 | F | childhood | GH | *MCPH1* | c.1561G>T<br>p.Glu521Ter<br>8-6302804-G-T | stop gained | 0 | CI | P | P | - |  |
| 43 | F | Young adult | GH | *SEC23B* | c.325G>A<br>p.Glu109Lys<br>20-18496339-G-A | missense | 0.00022 | P | LP | P | LP (0.994) |  |
| **Deleterious: AION Predictor and AI tools** |  |  |  |  |  |  |  |  |  |  |  |  |
| 1 | M | childhood | GH | *TMEM127* | c.380G>A<br>p.Arg127His<br>2-96920600-C-T | missense | 0 | US | VUS | P | LP (0.959) |  |
| 2 | M | adolescent | GH | *MSR1* | c.-4-1G>A<br>-<br>8-16035502-C-T | splice acceptor | 0.00085 | N/A | VUS | P | - | 1.0 (AL)<br>/99.7% |
| 3 | M | adolescent | GH | *BUB1* | c.2T>C<br>p.Met1? | start lost | 0 | N/A |  |  |  |  |
|  |  |  |  |  | 2-111435571-A-G |  |  |  |  |  |  |  |
| 10 | M | adolescent | GH | *POLE* | c.1274A>G<br>p.Lys425Arg<br>12-133250246-T-C | missense | 0.00002 | US | VUS | P | LP (0.84) |  |
| 10 | M | adolescent | GH | *SDHA* | c.1334C>T<br>p.Ser445Leu<br>5-236616-C-T | missense | 0 | CI | VUS | P | LP (0.9669) |  |
| 12 | M | childhood | PR L | *WRN* | c.2114C>T<br>p.Thr705Ile<br>8-30969156-C-T | missense | 0.00033 | US | VUS | LP | LP (0.9089) |  |
| 16 | F | adolescent | GH | *SDHD* | c.436G>A<br>p.Asp146Asn<br>11-111965650-G-A | missense | 0 | US | VUS | LP | LP (0.777) |  |
| 18 | F | childhood | GH | *RAD51C* | c.1026+6T>A<br>-<br>17-56809911-T-A | splice donor | 0 | CI | VUS | LB | - | 0.87 (DL)/<br>91.7% |
| 28 | F | Young adult | GH | *BAP1* | c.572T>C<br>p.Ile191Thr<br>3-52441198-A-G | missense | 0.00004 | CI | VUS | LP | LP (0.948) |  |
| 31 | M | Young adult | PR L | *POLG* | c.2641C>T<br>p.Pro881Ser<br>15-89864449-G-A | missense | 0.00004 | US | VUS | P | LP (0.917) |  |
| 48 | M | Young adult | GH | *POLG* | c.2615G>A<br>p.Ser872Asn<br>15-89864475-C-T | missense | 0 | N/A | VUS | P | LP (0.981) |  |
| **Ambiguous** |  |  |  |  |  |  |  |  |  |  |  |  |
| 17 | F | childhood | PR L | *SDHA* | c.1396G>A<br>p.Ala466Thr<br>5-236678-G-A | missense | 0.00086 | CI | VUS | P | A (0.4472) |  |
| 22 | F | Young adult | PR L | *PDE11A* | c.919C>T<br>p.Arg307Ter | stop gained | 0.00301 | CI | CI | P |  |  |
|  |  |  |  |  | 2-178879181-G-A |  |  |  |  |  |  |  |
| 44 | M | adolescent | PRL | TSC2 | c.5413G>C<br>p.Glu1805Gln<br>16-2138600-G-C | missense | 0 | US | VUS | LP | A (0.506) |  |

Overall, the deleterious germline variants involved the following genes: *BAP1, BRCA1, BUB1, ELAC2, FLCN, MCPH1, MSR1, MUTYH, PDE11A, POLE, POLG* (n=2)*, RAD51C, RECQL4, SDHA, SDHD, SEC23B, TMEM127* and *WRN* (Table 2). All but three subjects had single variants, while two subjects had two variants each.

### P/LP variants on ACMG and Clinvar

A pathogenic *BRCA1* variant was identified in a male subject with acrogigantism due to a somatotropinoma diagnosed during adolescence. While *BRCA1* is an established risk gene for breast/ovarian and other cancers, there is only one previous case report on *BRCA1* in a subject with a pituitary tumor (65).

A stop-gain P/LP variant in the *RECQL4* gene was identified in an adolescent male with acrogigantism. This DNA helicase gene is involved in DNA double stranded break repair, nucleotide excision repair, and base excision repair. Biallelic variants are associated with autosomal recessive developmental abnormalities and osteosarcoma risk (66). Recent studies have implicated heterozygous germline *RECQL4* variants with increased risk of ovarian and prostate cancer (67,68).

P/LP variants involving *FLCN* (frameshift) and *WRN* (missense) were noted in male pediatric subject that developed a giant prolactinoma (70 mm diameter at diagnosis). Germline *FLCN* variants are associated with renal tumors in sporadic populations and in those with the multisystem disease, Birt-Hogg-Dubé syndrome (69). Other potential neoplastic risks associated with germline *FLCN* variants include adrenal tumors, and one pituitary adenoma case was reported (70,71). This subject also had a missense germline *WRN* VUS that was classified as P/LP on AION predictor/Alpha Missense. *WRN* encodes a DNA helicase/exonuclease involved in DNA replication/repair, and germline variants in this gene lead to Werner syndrome, which is associated with progeria and increased cancer risk (72). *WRN* variants have also been identified in the setting of familial non-medullary thyroid cancer (73). A young female with a GH-secreting tumor had a variant in the DNA damage repair gene *MCPH1* and a concomitant VUS in the cancer risk gene *RAD51C.* A *SEC23B* variant was identified in a young adult female with acromegaly; this missense variant was judged pathogenic by ClinVar and AION predictor, likely pathogenic by ACMG, and Alpha Missense returned a high pathogenicity score (0.994). Germline heterozygous *SEC23B* variants have been identified at increased frequency in thyroid cancer and forms of Cowden syndrome (74).

A splice site variant in *ELAC2*, a prostate cancer risk gene, was found in a female subject with early-onset acromegaly; this was rated as P/LP by ACMG and AION predictor, and while the SpliceAI score (0.69) was borderline, the heuristic model from Sullivan *et* al scored it as 99.7% SAV. We also identified a P/LP *MUTYH* splice-site variant in an adolescent subject with acrogigantism due to a GH-secreting pituitary macroadenoma. Biallelic inactivating *MUTYH* variants lead to *MUTYH*-associated polyposis (MAP), that is associated with an increased risk of colorectal cancer. Heterozygous *MUTYH* variants have been linked to moderately-increased cancer risk in some settings (75).

### Deleterious variants using AION predictor and other tools

Some genes were previously implicated in the etiology of pituitary and other neuroendocrine tumors. One male with acrogigantism had a missense *SDHA* variant and a concomitant missense variant in *POLE* affecting the exonuclease domain of polymerase epsilon. Pathogenic variants in *SDHA, SDHD* and *TMEM127* have been identified in the paraganglioma-pheochromocytoma-pituitary adenoma association (3PA) (35,76). Germline *POLE* variants are associated with a familial risk of colorectal and other tumors (77).

Other variants affected genes with well-established tumor risk profiles. These included *BRCA-1 associated protein 1* (*BAP1)* (mesothelioma, melanoma, renal cell cancer risk), *BUB1* (colorectal cancer risk), and *RAD51C* (ovarian, other cancer risks) (78–80). Two male subjects, one with acrogigantism and another with a macroprolactinoma had variants in *POLG*. *POLG* encodes the gamma subunit of mitochondrial DNA polymerase. Rare disorders associated with loss of function of *POLG* include neuromuscular and metabolic diseases in children and adults (81). Germline variants in *POLG* have also been linked to several different cancers, such as, second neoplasms in pediatric cancer patients, and those with mesenchymal tumors or prostate cancer (82,83).

One subject with adolescent-onset acrogigantism had a deleterious variant in *MSR1* that was predicted to lead to loss of a splice acceptor site. *MSR1* encodes for a macrophage scavenger receptor, and pathogenic variants in *MSR1* have an established role in several cancer presentations, including hereditary gastric cancer, esophageal adenocarcinoma and prostate cancer (84–86).

### Ambiguous variants

Three subjects with prolactinomas had P/LP scores on AION predictor but VUS/conflicting significance scores on ACMG/ClinVar, ambiguous scores on Alpha Missense, or had a high prevalence in GnomAD. These were not included as potential P/LP variants. One *SDHA* variant occurred in a female pediatric subject with a macroadenoma; *SDHx* variants are associated with rare instances of pituitary tumors. One missense *TSC2* variant was identified in a teenage male with a macroprolactinoma. *TSC2* variants cause tuberous sclerosis, which has an increased risk of renal tumors and a handful of pituitary adenomas have been reported in tuberous sclerosis patients (87). A female with a macroprolactinoma had a frequently-identified heterozygous truncating variant in the *PDE11A* gene, which has previously been implicated in adrenal tumors, including adrenocortical carcinomas, and potentially pituitary adenomas (88–90).

### Copy number variations (CNV)

Germline DNA was also assessed for duplications and deletions affecting genes related to pituitary, neuro-endocrine and general cancer risk. One subject with acrogigantism due to a 65 mm GH-secreting macroadenoma diagnosed as a teenager had a small deletion on chromosome 7p22.1. This deletion included the DNA mismatch repair gene, *PMS2,* and was judged as pathogenic by ACMG.

### Pathway analysis

The gene ontogeny term enrichment analysis among the genes with P/LP variants is shown in Figure 1A. The top ranked significantly enriched heading was for DNA repair pathways-full network (WP4946; logP: -7.9). DisGeNET database assessment identified Hereditary Neoplastic Syndromes (GO term: C0027672; logP: -21.0) as the highest ranked disease term that was associated with the genetic variants identified in the young pituitary adenoma patients in the study.

**Figure 1.**
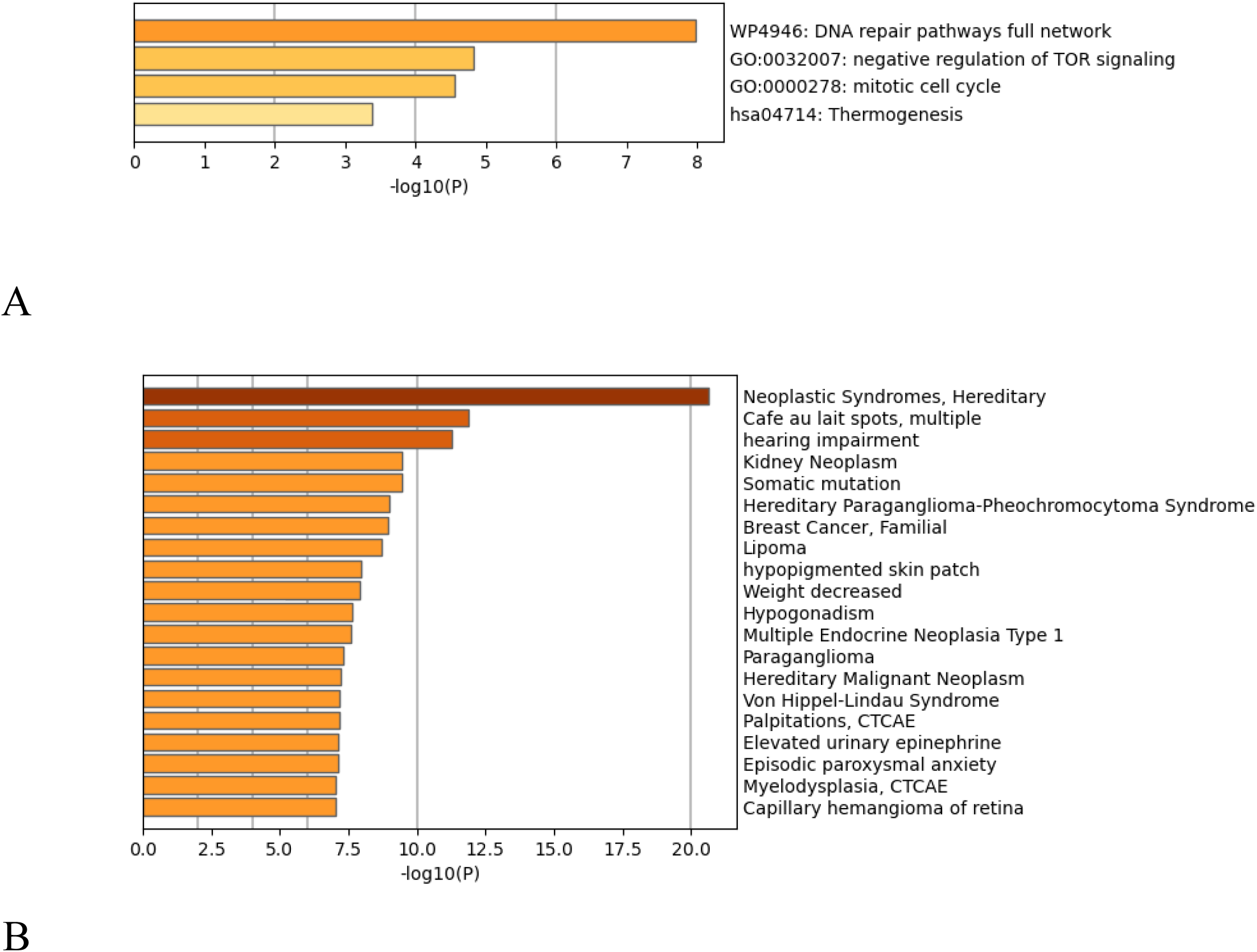
Metascape® representations of (**A**) enriched ontogeny terms and (**B**) disease-related DISGENET associations related to germline pathogenic/likely pathogenic variants in cancer-related genes identified in subjects with young-onset pituitary macroadenomas.

## Discussion

Deleterious germline variants are a rare but established cause of isolated anterior pituitary tumors, mainly restricted to clinical subgroups like pediatric patients. Even accounting for established risk genes like *AIP*, the etiology of most young-onset pituitary tumors remains obscure. In the current study we addressed this issue by performing WES analysis in a cohort with pituitary macroadenomas occurring before the age of 30, that were negative for known genetic causes. Using a comprehensive cancer gene target list of >200 genes, we identified deleterious germline sequence variants in 14.6%-31.3% of subjects, depending on the classification methodology. Apart from *SDHA* and *SDHD* none of the identified genes has been reliably implicated in pituitary adenomas. These results suggest that a wider range of inheritable tumorigenic pathways might contribute to pituitary tumorigenesis in children and young adults, particularly those with large somatotropinomas and prolactinomas.

Most germline genetic studies to date have employed multi-gene panels comprised of known endocrine/neuroendocrine tumor genes. Even in these, deleterious variants in *AIP, CDKN1B*, *MEN1*, *SDHx* and related genes are rarely identified (91–95). More recently, expanded gene sets that include some DNA mismatch repair genes have been deployed in pituitary adenoma populations. Using WES in a Saudi Arabian cohort of 134 non-familial pituitary adenomas, Alzahrani *et al* found germline P/LP variants in 6.7% of their cohort, including *AIP*, *CDH23*, *SDHA*, *DICER1*, *USP48*, *MSH2*, and *MLH1* (52). A Portuguese study of isolated sporadic macroadenoma subjects aged <40 years recently identified P/LP variants in 16/225 individuals (7.1%), although 6/16 variants affected the established pituitary tumor risk genes *AIP/MEN1* (53). As the Saudi cohort, that group used a limited panel of predominantly endocrine tumor-related genes (n=29). The Portuguese cohort was older by a decade than our population, and the Saudi study was performed in an older and milder population (>50% over 30 years at diagnosis; microadenomas included) than our study. By focusing on an extensively prescreened population using a comprehensive cancer gene panel, we expand the range of deleterious germline variants that may contribute to previously unexplained pituitary macroadenoma etiology in children and young adults.

While other groups have studied germline sequence variants, we also examined CNV affecting the cancer risk genes of interest. One subject had a small *PMS2* deletion, which would affect an established DNA mismatch repair gene that causes Lynch syndrome. Senter *et al* reported two pituitary adenomas in a large database analysis of pathogenic *PMS2* variant Lynch syndrome kindreds (96). In a Swedish national cohort of patients with Lynch syndrome due to mismatch repair gene pathogenic variants, Bengtsson *et al* reported three subjects with pituitary tumors, one of whom had a *PMS2* variant and a non-functioning pituitary adenoma (46). As noted above, the recent Portuguese report also identified subjects with germline *PMS2* variants (53). In contrast, we did not identify any sequence or copy number variants in other classical Lynch syndrome-associated genes (46,52,53,97,98).

Pituitary gigantism is a rare manifestation of acromegaly that is usually caused by a somatotropinoma during childhood or adolescence. Unlike acromegaly in general, the genetic pathophysiology of pituitary gigantism is known in about 50% of cases (99). In the current cohort that was negative for *AIP*/*CDKN1B/MEN1* variants/deletions and *GPR101* duplications, nine of the 14 gigantism subjects (64.3%) had sequence variants in *BRCA1, BUB1, MSR1, MUTYH, POLE/SDHA, RAD51C, RECQL4, POLG* or *SDHD*. These results further underline the importance of germline genetic factors in pituitary gigantism and suggests that the number of molecular pathways involved might be wider than previously thought.

Recent years have seen a re-assessment of the role of P/LP variants in the etiology of various cancers driven by widespread use of clinical exome/genome sequencing. Large studies have shown that universal germline sequencing in cancer patients increases the yield of P/LP variants markedly over that obtained with a stricter guideline focused sequencing approach (100). For example, Samadder *et al* found P/LP variants to be present in up to 12.5% with solid tumors treated in the Mayo Clinic group; importantly, only young age at cancer diagnosis was significantly predictive for identification of a genetic variant (101). Findings in common solid tumors have also been extended to the rarer, neuroendocrine tumor space. Perez and colleagues studied small bowel neuroendocrine neoplasms and 9-11% of subjects had a P/LP variant (43). Furthermore, Riechelmann *et al* reported that P/LP variant were present in nearly 16% of 108 subjects with gut or lung neuroendocrine neoplasms that occurred at a young age (18-50 years) (42). The most frequently implicated genes in that study were those related to DNA repair, as found in our cohort. Mohindroo *et al* had similar findings in a pancreatic NET population derived from two other major reference centers in the United States. Among 132 subjects with high-risk profiles (young age, personal/family history of cancer, and syndromic disease), P/LP variants were found in 33% of cases, most frequently *MEN1* or DNA repair pathway genes. In a validation cohort of 106 unselected pancreatic NET patients, 21% had a P/LP variant (41).

In contrast to the cancer studies mentioned above, pituitary lesions in our cohort were universally benign This raises a question as to whether germline genetic variants driving malignancy in other tissues can be of relevance for benign pituitary neoplasia. The example of syndromic conditions like MEN1 is instructive in this regard. The same *MEN1* germline pathogenic variant acting on the regulation of cell behavior across multiple tissue subtypes can lead to benign tumors in certain tissues (anterior pituitary, parathyroid) and malignant tumors (pancreatic NETs) in others (102). This is also seen in endocrine tumor syndromes with lower penetrance than MEN1 (35,103). Following a similar model, it is possible that deleterious variants in DNA repair genes typically associated with hereditary breast/ovarian, or colorectal cancers could also manifest as pathologically benign adenomas in the anterior pituitary. The epidemiology of pituitary adenomas/PitNETs is relevant. Pituitary tumors are often found incidentally as tiny innocuous lesions in imaging and pathology studies (prevalence of about 1:5), whereas very few progress to cause clinically apparent disease (prevalence of 1:1000) (104). Only a small minority of pituitary tumors progress to aggressive or locally invasive tumors and vanishingly few become carcinomas. Due to its extreme importance regulating multiple vital pathways, the anterior pituitary may be a molecularly privileged tissue that is relatively protected against malignant transformation. Somatic events play an important role in removing the brakes on neoplastic proliferation, as activating *GNAS* and *USP8* are frequent causes of acromegaly and Cushing’s disease, respectively, but do not seem to cause aggressive disease (10,105,106). Although germline factors like *AIP* can lead to tumors with an unfavorable clinical phenotype, these almost never undergo malignant transformation (25). Aggressive pituitary adenomas and pituitary carcinomas are among the most challenging patients to manage therapeutically (107–110). Somatic DNA studies of resected pituitary adenoma tissue have revealed recurrent pathogenic variants in genes such as *ATRX* and *TP53*, in patients with aggressive corticotroph tumors or carcinomas (111–114). Germline studies in these aggressive adenomas and carcinomas have been limited to case reports and small series, although cases involving *CHEK2* support a role for “traditional” cancer genes in pituitary adenoma etiology (49,51,115). Taken together we suggest that deleterious germline variants in cancer-risk genes play a more important role that considered to date in the etiology of pituitary adenomas/PitNETs.

Guidelines for acromegaly and other pituitary tumor subtypes do not yet include specific management recommendations regarding genetic testing, possibly due to the relative rarity of established germline genetic causes overall (<5% of cases) (22). The 2022 WHO classification of pituitary tumors as PitNETs focuses by necessity on surgically resected tissues and does not yet extend to outcomes in pre-surgical patients (2,3,116,117). Recently, Ho and colleagues have proposed a novel scoring system that incorporates multiple clinically relevant features of pituitary adenomas at presentation and during treatment (1). This system introduces germline genetic variants in *AIP*, *MEN1* and other genes into predicting disease severity (1). Such a systematic approach to classify pituitary tumor patients could be expanded to include newer genetic factors, once these have been established and validated.

Our study has several limitations in terms of scope and interpretation. Ideally, a study would have access to paired germline and tumor DNA to assess whether a second hit in an affected gene is present and to describe the somatic pattern of chromosomal disturbances that are characteristic of pituitary adenomas (15–18,21). A major challenge in clinical genetics is the interpretation of rare variants (particularly missense and splice variants) that lack definitive functional studies to determine pathogenicity. For this reason, *in silico* tools to predict functional effects have proliferated and have become increasingly sophisticated when combined into compendium models or AI-assisted toolsets like we used here. However, no model can reliably replace functional assays, and there remains a risk for the classification of potentially innocuous variants as deleterious, thereby increasing reported rates. In this study we purposely separated more traditional classification criteria from novel *in silico* and AI driven models to acknowledge this uncertainty about classification of cancer gene variants in pituitary tumor patients. Furthermore, the mechanisms by which these putative deleterious variants could influence pituitary neoplastic transformation remain to be studied and established. Also, the patient population does not reflect the general epidemiological profile of pituitary adenomas in the young, which are more often small prolactinomas, patients with Cushing’s disease or those with non-functioning adenomas (104). This skewing comes from the recruitment criteria of our ongoing research studies into pituitary gigantism and aggressive/large pituitary adenomas, leading to a relative over-representation of somatotropinomas.

When taken as a whole, results to date suggest that deleterious germline variants in cancer risk genes play a more important role in pituitary tumor pathogenesis than was previously thought. In line with similar studies in neuroendocrine and other cancers, pituitary adenomas may be a clinical manifestation of underlying germline cancer risk alleles. In the pediatric-adolescent cohort of pituitary gigantism patients, the high prevalence of deleterious germline variants confirms that somatotropinomas in the young appear to be particularly sensitive to germline genetic pathology. In conclusion, the germline genetics of pituitary adenomas in young subjects appears to mirror that of neoplasia in many other tissues. More widespread use of exome/genome sequencing may allow for the identification of deleterious variants that can permit improved family screening or even identify novel druggable pathways for expanded personalized therapy.

## Data Availability

All data produced in the present study are available upon reasonable request to the authors

## Author contributions

Adrian F. Daly: Conceptualization, Methodology, Resources, Investigation, Formal Analysis,

Writing-Original Draft, Writing-Review and Editing

Marie-Lise Jaffrain-Rea: Resources, Investigation, Writing-Review and Editing

Kalyani Sridharan: Resources, Investigation, Writing-Review and Editing

Giampaolo Trivellin: Methodology, Validation, Writing-Review and Editing

Francesca Carbonara: Resources, Investigation, Writing-Review and Editing

Wouter De Herder: Resources, Investigation, Writing-Review and Editing

Ismene Bilbao: Resources, Investigation, Writing-Review and Editing

Maria Segni: Resources, Investigation, Writing-Review and Editing

Margaret Zacharin: Resources, Investigation, Writing-Review and Editing

Maryna Solovey: Resources, Investigation, Writing-Review and Editing

Ulrich Paetow: Resources, Investigation, Writing-Review and Editing

Nalini Shah: Resources, Investigation, Writing-Review and Editing

Tushar Bandgar: Resources, Investigation, Writing-Review and Editing

Liliya Rostomyan: Resources, Investigation, Writing-Review and Editing

Sebastian J.C.M.M. Neggers: Resources, Investigation, Writing-Review and Editing

Albert Beckers: Resources, Investigation, Supervision, Writing-Review and Editing, Funding Acquisition

Patrick Pétrossians: Methodology, Software, Validation, Data Curation, Formal Analysis, Supervision, Writing-Review and Editing, Funding Acquisition

## Acknowledgements

The authors would like to acknowledge Dr. Aina Pi Roig and Dr. Rocio Acuña of Nostos Genomics GmbH for their advice and assistance on bioinformatics, data handling and discussions on AION predictor. We thank all the supporting clinicians for contributing genetic material and clinical details.

