## Supplemental File 1 for "High prevalence of deleterious germline variants in cancer risk genes among subjects with young-onset, sporadic pituitary macroadenomas"

**Methods (see Supplemental Figure 1)**

VCF files initially underwent comprehensive QC to verify upstream processes such as sequencing and bioinformatics analysis and includes consistency checks on VCF file structure and content, verification of submitted information as well as detection and correction of file and data variations to adhere to best practices for germline variant calling in clinical sequencing (1). Variant annotation is carried out on the filtered variants using AION predictor's annotation engine, based on Ensembl's Variant Effect Predictor (VEP) enhanced with additional functionality developed in-house. Variants are annotated with >60 data properties pertaining to predicted and observed molecular effects, variation observed in healthy population and in individuals with genetic disorders, evolutionary conservation, clinical phenotypes to provide comprehensive molecular, clinical and population frequency annotations. Annotation data is extracted from multiple data sources including external databases (ENSEMBL, RefSeq, UniProt, UCSC, gnomAD, ClinVar, ClinGen, GenCC, HPOjax, Monarch, Orphanet, and published scientific literature) and internal knowledge bases (e.g. variant frequencies from the user's internal cohort, previous classifications, etc.). Variant annotation and further analysis were performed using the GRCh37 reference.

Following recommendations by Austin-Tse *et al* concerning best practices for variant interpretation, all variants analyzed were annotated with ClinVar classifications (including known pathogenic variants in non-coding regions outside of coding sequence and flanking intronic regions) (2). AION predictor includes ClinVar data with additional pre-processing to increase sensitivity for pathogenic variants by solving variants with conflicting interpretations. Variant classification was carried out on all filtered variants in coding regions (+/- 50 bp intronic regions) through an automated implementation of the standards and guidelines from the American College of Medical Genetics (ACMG) (3).

To detect variants currently classified as variants of uncertain significance, but with molecular characteristics indicating high likelihood of being pathogenic, all filtered variants were classified with AION predictor's proprietary classification algorithm. This algorithm is a probabilistic graphical model which analyzes the molecular properties of individual genetic variants to classify them as "Pathogenic" or "Benign". The classifications produced by the model are accompanied by a posterior probability of the variant being pathogenic, ranging from 0 (0% probability of the variant being pathogenic) to 1 (100% probability of the variants being pathogenic). Additionally, the variant classification model produced intermediate predictions of probability of functional disruption at a molecular level accompanied by a posterior probability, with predictions of variant effects at the RNA and protein level, as well as predictions of constraint to variation in the affected gene and genomic region. This approach detected pathogenic mutations among variants of unknown significance with high sensitivity, by providing prospective variant classifications even in the absence of sufficient evidence to support a definitive classification. All variants classified as “Pathogenic” or “Likely pathogenic” by at least one of these three classification methods were considered for variant prioritization. AION predictor’s proprietary prioritization algorithm analyzed genetic and molecular data from all candidate pathogenic variants identified together with case clinical data to produce a ranked list of combinations of candidate pathogenic variant(s) and associated disease(s).

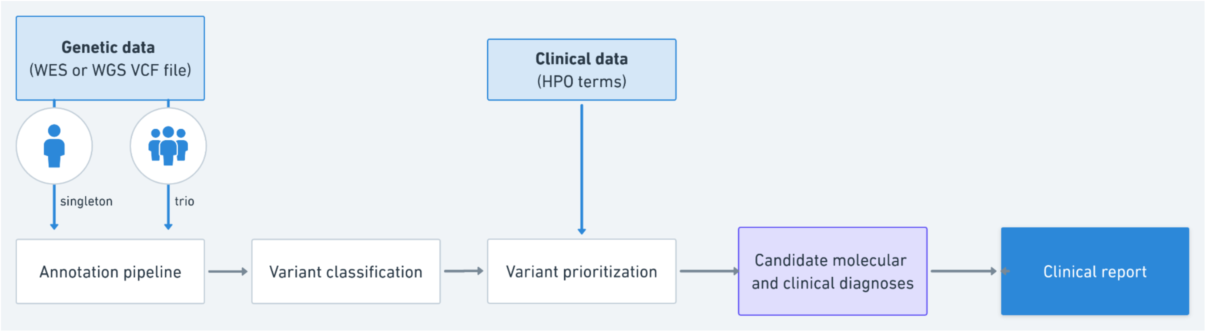


**Supplemental Figure 1: AION predictor genetic variant analysis workflow**

Clinical features observed in the patient undergoing genetic testing were provided as Human Phenotype Ontology (HPO) terms, providing a standardized description of phenotypic abnormalities related to human genetic diseases. For the current study, the HPO terms were restricted to “pituitary adenoma” and specific pituitary adenoma sub-types (e.g. “Pituitary growth hormone cell adenoma”, etc.). Clinical feature overlap was calculated for all diseases associated with genes in which candidate disease-causing variants were identified. Phenotype overlap was calculated using the patient and disease clinical features, input as HPO terms with term propagation. Variant characteristics indicating severity of variant effects, such as predicted functional impact and variant frequency in large population databases, were assessed during variant prioritization. Variants with high severity predicted consequences (e.g. LOF mutations or gene deletions) and variants that are rare or absent from population databases (e.g. gnomAD) are ranked as higher priority candidates.

Following this process above, a soft filter was applied to restrict variants to a comprehensive list of genes that are established/associated with a risk of solid cancers. The full listing was derived from internal AION predictor gene filters, supplemented by literature-based resources and included novel pituitary adenoma associated genes (3–10).

*ACD, AIP, ALK, AP2S1, APC, APOBEC3B, AR, ARMC5, ASPM, ATM, ATR, ATRX, AXIN2, BAP1, BARD1, BLM, BMPR1A, BRAF, BRCA1, BRCA2, BRIP1, BUB1, BUB1B, CABLES1, CASR, CBL, CDC73, CDH1, CDH10, CDH23, CDK4, CDKN1B, CDKN1C, CDKN2A, CDKN2B, CEP57, CFAP100, CHEK2, CTC1, CTNNA1, CTR9, CXCR4, CYLD, DDB2, DICER1, DIRAS3, DIS3L2, DKC1, DLST, DNMT3, EGFR, EGLN1, ELAC2, ELP1, EPAS1, EPCAM, ERBB4, ERCC1, ERCC2, ERCC3, ERCC4, ERCC5, ESR1, EXT1, EXT2, EZH2, FADD, FANCA, FANCB, FANCC, FANCD2, FANCE, FANCF, FANCG, FANCI, FANCL, FANCM, FAT1, FBXW7, FEN1, FH, FLCN, FOXE1, GCGR, GCM2, GNA11, GNAS, GPC3, GPR101, GPR161, GREM1, HABP2, HGF, HNF1A, HOXB13, HRAS, KDR, KIF1B, KISS1R, KIT, KRAS, LMO1, LRP5, LRP6, LZTR1, MAD2L2, MAP2K1, MAP2K2, MAX, MBD4, MCPH1, MDH2, MEN1, MET, MINPP1, MITF, MLH1, MPL, MSH2, MSH3, MSH5, MSH6, MSR1, MUC6, MUTYH, NBN, NDUFA13, NF1, NF2, NHP2, NKX2-1, NOP10, NRAS, NSD1, NTHL1, NYNRIN, PALB2, PAM, PARN, PAX5, PDE11A, PDGFRA, PHOX2B, PIK3CA, PKD1, PMS1, PMS2, POLD1, POLE, POLG, POLH, POLQ, POT1, PPM1D, PPP1CB, PRF1, PRKAR1A, PRLR, PTCH1, PTCH2, PTCSC1, PTCSC3, PTEN, PTPN11, PTPN13, RAD50, RAD51C, RAD51D, RAD54L, RAF1, RB1, RECQL4, REST, RET, RFWD3, RHBDF2, RIT1, RNASEL, RNF43, RPS20, RRAS2, RTEL1, SBDS, SDHA, SDHAF2, SDHB, SDHC, SDHD, SEC23B, SEMA4A, SETBP1, SHOC2, SLC25A11, SLC5A5, SLX4, SMAD4, SMARCA4, SMARCB1, SMARCE1, SOS1, SOS2, SPOP, SQSTM1, SRGAP1, SRRM2, STAT3, STK11, SUCLG2, SUFU, TERC, TERF2IP, TERT, TG, TGFBR2, TINF2, TMEM127, TP53, TP63, TRIM28, TRIM37, TRIP13, TSC1, TSC2, TSHR, UBE2T, USP8, USP9X, VHL, VTRNA2-1, WAS, WRAP53, WRN, WT1, XPA, XPC, XRCC2*

**Supplemental Table 1: Germline cancer risk gene panel**
